# Clinical Risk Factors for COVID-19 related Severe Outcome

**DOI:** 10.1101/2021.11.30.21267086

**Authors:** Chaorui C. Huang

## Abstract

**Background:** We aimed to evaluate the risk factors for Coronavirus disease 2019 (COVID-19) related severe outcome in New York State (NYS) and proposed a method that could be used to inform future work to develop clinical algorithms and predict resource needs for COVID-19 patients.

**Methods:** We analyzed COVID-19 related hospital encounter and hospitalization in NYS from April 1^st^ to November 17^th^, 2020, using Statewide Planning and Research Cooperative System (SPARCS) hospital discharge dataset. Logistic regression was performed to evaluate the risk factors for COVID-19 related in-hospital death using demographic variables, symptom, rapid clinical examination, and medical history of chronic co-morbid conditions. Receiver operating characteristic (ROC) curve was calculated, and cut-off points for predictors were selected to stage the risk of COVID-19 related fatal outcome.

**Findings:** Logistic regression analysis showed age was the greatest risk factor for COVID-19 related fatal outcome, which by itself achieved the diagnostic accuracy of 0.78 represented by the area under the ROC curve. By adding other demographic variables, dyspnea or hypoxemia and multiple chronic co-morbid conditions, the diagnostic accuracy was improved to 0.85. We selected cut-off points for predictors and provided a general recommendation to categorize the levels of risk for COVID-19 related fatal outcome.

**Interpretation:** We assessed risk factors associated with in-hospital COVID-19 mortality and identified cut-off points that might be used to categorize the level of risk. Further studies are warranted to evaluate laboratory tests and develop laboratory biomarkers to improve the diagnostic accuracy for early intervention.

## Introduction

Given the heterogeneous clinical presentation and outcomes of Coronavirus disease 2019 (COVID-19), and the scope of the outbreak, there is an urgent need to develop a classified system for clinical diagnosis, which can identify the at-risk patients for early intervention, and effectively flag and track patients who are in need, as well as accurately calculate the medical supplies, and properly allocate resources and staffs for outbreak response.^1^

There has been a list of risk factors developed for the COVID-19 related severe outcomes by the on-going research.^2^ However, it wasn’t clear how the multiple risk factors with different combinations weigh in a single person, and exactly who should be prioritized for early intervention. Without such knowledge, it is also difficult to calculate the daily medical supply. The window for early intervention is shortly closed, and a measurement that can rapidly evaluate and screen the at-risk patients for severe outcome is in need.

Another concern is that most of the current on-going epidemiology studies primarily focused on the population-level results, such as population probabilities. However, the results are hard to interpret in clinical settings. Specifically, speaking of the probability of developing severe outcome, an individual patient can only have two possible outcomes, which is either they progress to a severe stage of disease or they don’t. The population-level results, which only provide the population probabilities, are difficult to use to facilitate targeted interventions for an individual patient. This applies to both clinical settings, while physicians treat patients, as well as to public health settings, when public health specialists investigate and follow-up individuals for surveillance purpose.

In this study, we aimed to evaluate the risk factors for COVID-19 related severe outcome, specifically in-hospital death in New York State (NYS) and propose a strategy, which can be directly applied to clinic to rapidly screen the at-risk patients for early intervention. It will also aid the daily clinical operation, such as medical supply calculation, as well as resource and staff allocation.

## Methods

### Data Source and Study Population

We analyzed Statewide Planning and Research Cooperative System (SPARCS) hospital discharge data for NYS residents (based on address of home residence) who were either hospitalized or visited ambulatory surgery or emergency department or outpatient (we referred them as hospital visit throughout the paper), because of COVID-19 from April 1^st^ to November 17^th^, 2020. We also conducted post-hoc analysis in two separated sub-samples in New York City (NYC), which included the five boroughs of Manhattan, Queens, Bronx, Brooklyn, and Staten Island, and in other NYS regions.

SPARCS is a comprehensive all payer data reporting system that collects discharge data from all hospitals in NYS.^3^ Each discharge record within SPARCS includes a principal diagnosis and up to 24 secondary diagnoses, coded using the *International Classification of Diseases, 10th revision, Clinical Modification* (*ICD-10-CM)*.^*4*^

### Definitions

We identified COVID-19 related hospitalizations and hospital visits by examining the principal diagnosis. Of these records, we further identified in-hospital death, which served as the main study outcome. We identified specific COVID-19 related clinical presentation/examination and chronic co-morbid conditions by examining the secondary diagnosis. The clinical presentation/examination and chronic co-morbid conditions included dyspnea or hypoxemia, overweight or obesity, essential (primary) hypertension, diabetes mellitus, hyperlipidemia, chronic cardiovascular disease, chronic kidney disease, chronic pulmonary disease, malignant neoplasms, dementia, human immunodeficiency virus (HIV), cerebral palsy, sickle-cell disorders, asthma, nicotine dependence, and pregnancy. The chronic co-morbid conditions were selected by manual review of the secondary diagnosis of COVID-19 related hospital encounter, as well as the risk profile provided by Centers for Disease Control and Prevention (CDC).^2^

### Data Analysis

The unit of analysis was a hospitalization or hospital visit at ambulatory surgery or emergency department or outpatient. We firstly conducted descriptive statistics and calculated in-hospital death, and length of hospital stay in NYS. We then performed logistic regression to evaluate the risk factors for the COVID-19 related severe outcome (in-hospital death) in NYS. We also repeated the regression analysis using the sub-sample of NYC and other NYS regions separately. Receiver operating characteristic (ROC) curve was calculated as an estimate of diagnostic accuracy for COVID-19 related in-hospital death.

Prior to building a logistic regression model, the association among categorical variables were examined by Chi-square test, and Cramer’s V was used to determine the strength of the association. It was well established that age was associated with multiple co-morbid conditions, such as hypertension, diabetes mellitus, cardiovascular disease, dementia.

We built up two logistic regression models, which were “age model” and “all effect model”. The outcome was COVID-19 related in-hospital death. The predictors were demographic variables (age, sex, race/ethnicity), clinical presentation/examination (dyspnea or hypoxemia), and chronic co-morbid conditions (overweight or obesity, essential (primary) hypertension, diabetes mellitus, hyperlipidemia, chronic cardiovascular disease, chronic kidney disease, chronic pulmonary disease, malignant neoplasms, dementia, HIV, cerebral palsy, sickle-cell disorders, asthma, nicotine dependence, and pregnancy). Of note, race/ethnicity itself is not a predictor for severe COVID-19 outcomes, but rather a proxy for unmeasured social context/factors, including structural vulnerability and racism.

For each model building, we followed the following steps. Step 1: Build preliminary main effects model, that a simple logistic regression was fitted for each variable. Variables with p-value <0.05 were candidates for multiple logistic regression model. In the multiple regression model, the variables were removed if the p-value was >0.05 or the change of odds ratio was less than 15% from 1, which was considered not clinically meaningful. Step 2: Scale checking for the linear variables was performed under the assumption that log odds of outcome (COVID-19 related in-hospital death) increased by the same fixed amount anywhere on the X scale (predictors). Step 3: Check for possible interactions. Step 4: We calculated ROC curve for each model. The diagnostic accuracy was considered optimal with 0.9-1.0 (or 90%-100%) area under ROC curve; acceptable with 0.8-0.9 (or 80%-90%) area under ROC curve; fair with 0.7-0.8 (or 70%-80%) area under ROC curve; and poor with 0.6-0.7 (or 60%-70%) area under ROC curve. The Brier score was also calculated. Step 5: We calculated the predicted odds and probability for the outcome among the individual subjects, and generated sensitivity and specificity table. Based on the sensitivity and specificity table, we selected the cut-off points of odds and probability, and provided a general recommendation to stage the risk of fatal outcome among the COVID-19 patients.^5^

All statistical analyses were performed using SAS, version 9.4, SAS Institute.

### Study Approval

This activity was determined by NYC Department of Health and Mental Hygiene (DOHMH) to involve the use of existing data and was exempt from DOHMH Institutional Review Board review.

## Results

### Descriptive Statistics

From April 1^st^ to November 17^th^ in 2020, there were 102,440 COVID-19 hospitalizations or visits at ambulatory surgery or emergency department or outpatient in total in NYS, among which 61,296 (59.8%) were from NYC, and 41,144 (40.2%) were from other regions in NYS. The overall COVID-19 related percentage of in-hospital death was 9.9% (NYC: 11.4%; other NYS regions: 7.5%). The overall COVID-19 related percentage of in-hospital death in NYS was 0.3% among children less than 18 years old (NYC: 0.3%; other NYS regions: 0.4%); 3.8% among adults from 18 to 65 years old (NYC: 4.5%; other NYS regions: 2.8%) and 20.9% among elderly older than 65 years old (NYC: 24.5%; other NYS regions: 15.8%). A total of 97.1% of the in-hospital death occurred among the hospitalized patients, and 2.9% at emergency department. The average length of hospital stay was 9.4±11.3 (standard deviation (SD)) days in NYS (NYC: 9.3±11.6 SD; other NYS regions: 9.5±10.6 SD).

### Logistic Regression Estimates in NYS

The results of maximum likelihood estimate of logistic regression and odds ratio for COVID-19 related severe outcome in NYS were shown in Table 1.

**Table 1.**
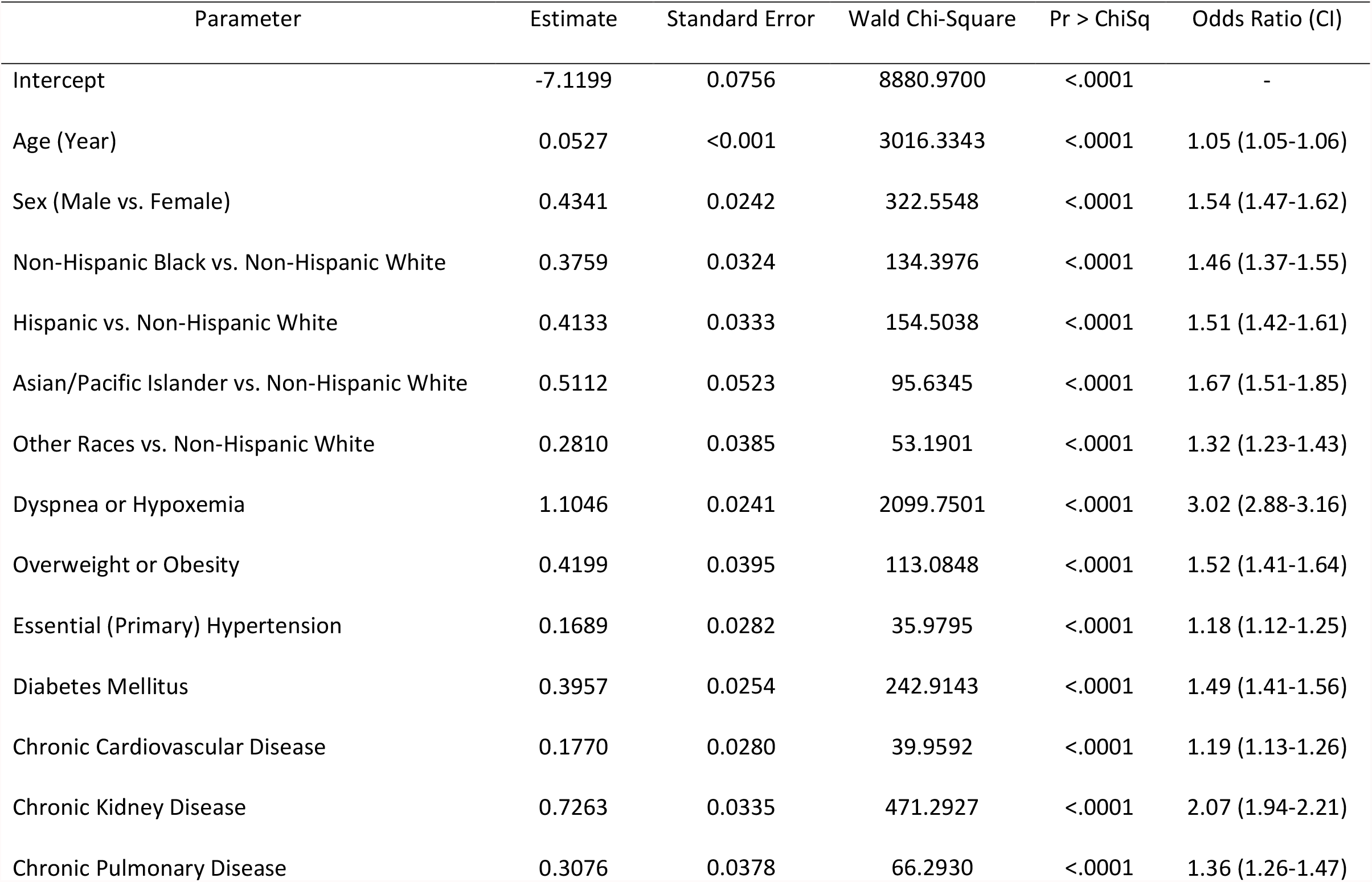
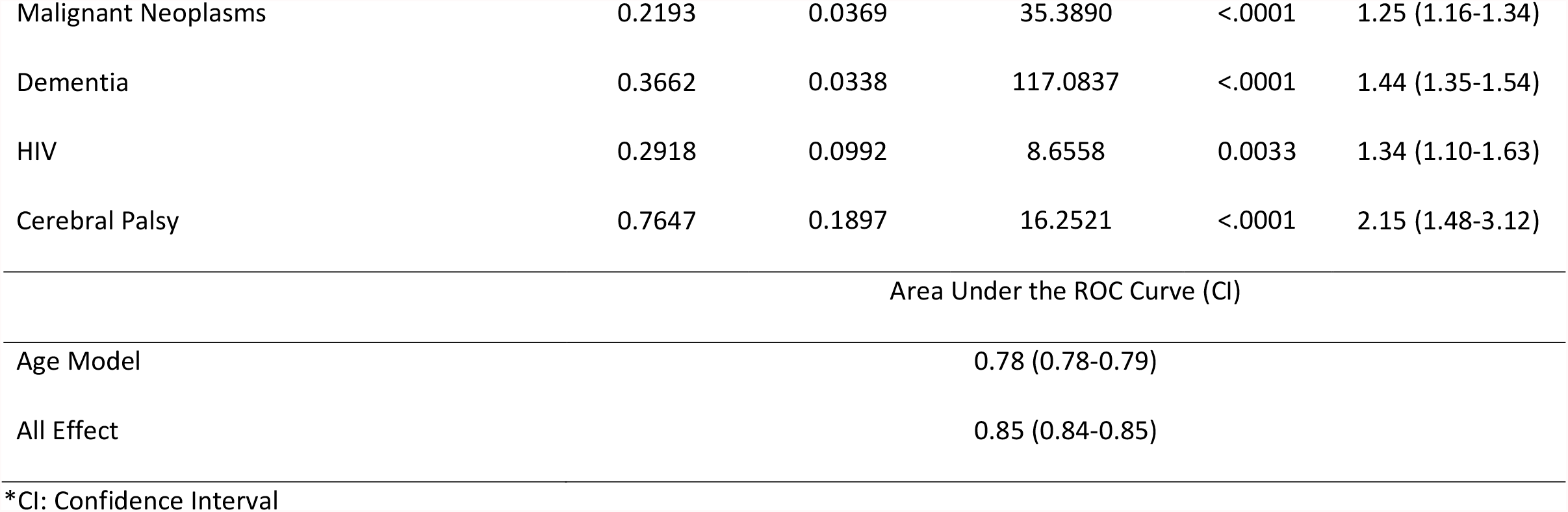
Logistic Regression Estimates of the Risk Factors of COVID-19 related in-Hospital Death in NYS.

| Parameter | Estimate | Standard Error | Wald Chi-Square | Pr > ChiSq | Odds Ratio (CI) |
| --- | --- | --- | --- | --- | --- |
| Intercept | -7.1199 | 0.0756 | 8880.9700 | <.0001 | - |
| Age (Year) | 0.0527 | <0.001 | 3016.3343 | <.0001 | 1.05 (1.05-1.06) |
| Sex (Male vs. Female) | 0.4341 | 0.0242 | 322.5548 | <.0001 | 1.54 (1.47-1.62) |
| Non-Hispanic Black vs. Non-Hispanic White | 0.3759 | 0.0324 | 134.3976 | <.0001 | 1.46 (1.37-1.55) |
| Hispanic vs. Non-Hispanic White | 0.4133 | 0.0333 | 154.5038 | <.0001 | 1.51 (1.42-1.61) |
| Asian/Pacific Islander vs. Non-Hispanic White | 0.5112 | 0.0523 | 95.6345 | <.0001 | 1.67 (1.51-1.85) |
| Other Races vs. Non-Hispanic White | 0.2810 | 0.0385 | 53.1901 | <.0001 | 1.32 (1.23-1.43) |
| Dyspnea or Hypoxemia | 1.1046 | 0.0241 | 2099.7501 | <.0001 | 3.02 (2.88-3.16) |
| Overweight or Obesity | 0.4199 | 0.0395 | 113.0848 | <.0001 | 1.52 (1.41-1.64) |
| Essential (Primary) Hypertension | 0.1689 | 0.0282 | 35.9795 | <.0001 | 1.18 (1.12-1.25) |
| Diabetes Mellitus | 0.3957 | 0.0254 | 242.9143 | <.0001 | 1.49 (1.41-1.56) |
| Chronic Cardiovascular Disease | 0.1770 | 0.0280 | 39.9592 | <.0001 | 1.19 (1.13-1.26) |
| Chronic Kidney Disease | 0.7263 | 0.0335 | 471.2927 | <.0001 | 2.07 (1.94-2.21) |
| Chronic Pulmonary Disease | 0.3076 | 0.0378 | 66.2930 | <.0001 | 1.36 (1.26-1.47) |
| Malignant Neoplasms | 0.2193 | 0.0369 | 35.3890 | <.0001 | 1.25 (1.16-1.34) |
| Dementia | 0.3662 | 0.0338 | 117.0837 | <.0001 | 1.44 (1.35-1.54) |
| HIV | 0.2918 | 0.0992 | 8.6558 | 0.0033 | 1.34 (1.10-1.63) |
| Cerebral Palsy | 0.7647 | 0.1897 | 16.2521 | <.0001 | 2.15 (1.48-3.12) |

Area Under the ROC Curve (CI)
|  |  |
| --- | --- |
| Age Model | 0.78 (0.78-0.79) |
| All Effect | 0.85 (0.84-0.85) |

\*CI: Confidence Interval

The first model with only age as a predictor showed that age was a significant risk factor for COVID-19 related in-hospital death. It achieved a diagnostic accuracy of 0.78, represented by the area under the ROC curve. (Table 1, Figure 1)

**Figure 1.**
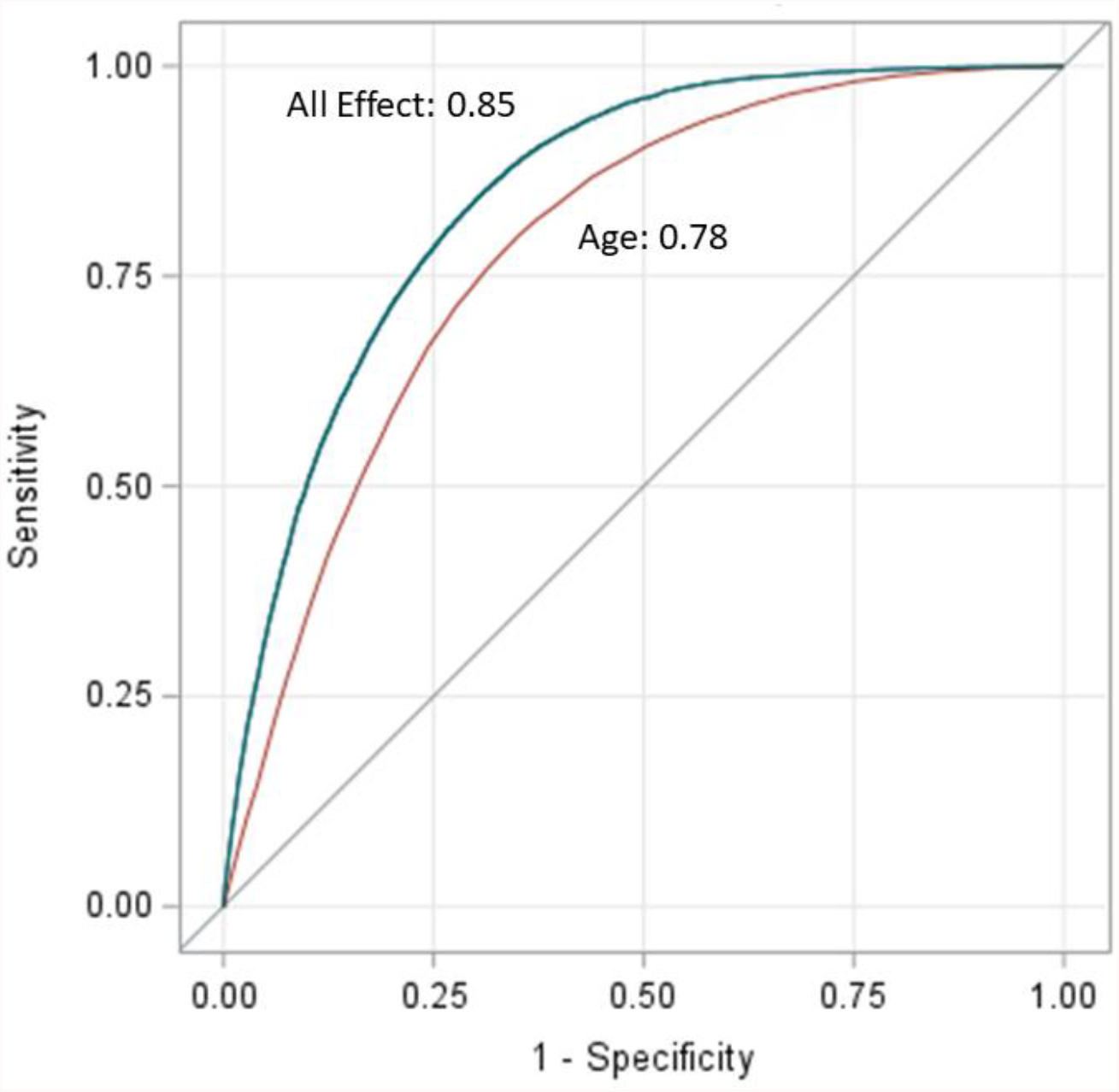
Receiver Operating Characteristic (ROC) Curve as an Estimate of Diagnostic Accuracy for COVID-19 related in-Hospital Death in NYS. The area under the ROC curve for age model was 0.78. The area under the ROC curve for combined effect of demographic variables (age, sex, race/ethnicity), clinical presentation/examination, and all chronic co-morbid conditions was 0.85.

In the second “all-effect” model, we added the covariates step-by-step. By including the chronic co-morbid conditions together with age, the diagnostic accuracy improved from 0.78 to 0.82. In the final model, demographic variables (age, sex, race/ethnicity), clinical presentation/examination (dyspnea or hypoxemia), and chronic co-morbid conditions (overweight or obesity, essential hypertension, diabetes mellitus, chronic cardiovascular disease, chronic kidney disease, chronic pulmonary disease, malignant neoplasms, dementia, cerebral palsy) were significant predictors for COVID-19 related in-hospital death. The diagnostic accuracy of this final model for predicting the COVID-19 related fatal outcome was 0.85, represented by the area under the ROC curve. The automated stepwise model selection didn’t include HIV as a risk factor in the final model. We still included HIV in the final model, because it was significant in the manual stepwise model selection, but considered that HIV was likely a borderline significant risk factor for COVID-19 related fatal outcome. (Table 1, Figure 1) The Brier score was 0.0741, which indicted a good predictive accuracy.

With further manual calculation based on the coefficient in Table 1 for “all-effect” model, the results showed that the odds of a COVID-19-related fatal outcome for 65 year-old patients was 11.9 times the odds of 18 year-old patients, and 23.6 times the odds of 5 year-old patients, after accounting for sex, race/ethnicity, dyspnea or hypoxemia and chronic co-morbid conditions. Patients of Asian ancestry had the highest odds for COVID-19 related fatal outcome among all races, after accounting for age, sex, dyspnea or hypoxemia, and chronic co-morbid conditions. The odds ratio of chronic co-morbid conditions for COVID-19 related fatal outcome typically ranged between 1.0-3.0, after correcting for demographic variables and dyspnea or hypoxemia. (Table 1 & 4)

We didn’t find sickle-cell disorders, asthma, or nicotine dependence to be significant in the final model in this dataset. There weren’t enough cases labeled with pregnancy status to make the model converge, so we were unable to estimate the risk of pregnancy. Hyperlipidemia had mild to moderate association with essential hypertension (Cramer’s V=0.32) and diabetes mellitus (Cramer’s V=0.36). We conducted an additional analysis by combining hyperlipidemia, essential hypertension, and diabetes mellitus, which resulted in a similar diagnostic accuracy of 0.85 with the area under the ROC curve. Considering the potential duplicated visits or clinical encounters, we conduced additional analysis removing the duplicated personal identifiers, but results only changed minimally.

### Risk Staging

The ROC curve, which evaluated how well a continuous predictor can classify a binary outcome, was plotted based on the sensitivity and specificity table. The cut-off points of the predictors (predicted odds and/or probability), which can provide the most optimal sensitivity and specificity for diagnostic classification, were evaluated. The ideal cut-off point is supposed to be the predictor value corresponding to the point on the ROC curve, which is closest to the upper left corner. It was easy to define such cut-off point when a diagnostic test provided both high sensitivity and specificity. However, in our study, with the moderate diagnostic accuracy of 0.85, we proposed two methods for cut-off point selection.

For method I, we chose the nearest point to the upper left corner of the ROC curve graph and classified the patients to high-risk group vs. low-risk group for the COVID-19 related mortality.^5^ For method II, we proposed a range of cut-off points and classified the risk of the COVID-19 related in-hospital death into five stages. We arbitrarily selected four cut-off points of odds and/or probability, which corresponded to the sensitivity and specificity level at 95% and 80% on the ROC curve, separately. Five levels of risk for COVID-19 related severe outcome were ranked, which were high risk for mortality, at-risk (high end) for mortality, at-risk for mortality, at-risk (low end) for mortality, and low risk for mortality. We also provided additional cut-off points of odds and/or probability and corresponding sensitivities and specificities in Table 2. Clinicians can choose to use different cut-off points based on their own clinic needs for i.e. diagnostic or supply calculation purpose. (Table 2 & 3)

**Table 2.** Selected Odds and Probability Cut-off Points for COVID-19 related Severe Outcome and Corresponding Sensitivity and Specificity.

| Odds | Probability | Sensitivity | Specificity |
| --- | --- | --- | --- |
| 1.068 | 0.517 | 9.4% | 99.0% |
| 0.474* | 0.321* | 32.1%* | 95.0%* |
| 0.286 | 0.222 | 50.6% | 90.0% |
| 0.200 | 0.167 | 62.3% | 85.0% |
| 0.149* | 0.129* | 71.6%* | 80.0%* |
| 0.106* | 0.096* | 80.0%* | 73.7%* |
| 0.101* | 0.092* | 81.2%* | 72.6%* |
| 0.085 | 0.078 | 85.0% | 69.0% |
| 0.064 | 0.060 | 90.0% | 62.9% |
| 0.042* | 0.040* | 95.0%* | 53.2%* |
| 0.018 | 0.018 | 99.0% | 32.2% |

\*Generally recommended cut-off points (see Table 3)

**Table 3.** General Recommendation for Staging the COVID-19 related Severe Outcome.

| Staging | Odds | Probability | Interpretation |
| --- | --- | --- | --- |
| Method I |  |  |  |
| Stage II | $\geq 0.101$ | $\geq 0.092$ | High risk for mortality |
| Stage I | $< 0.101$ | $< 0.092$ | Low risk for mortality |
| Method II |  |  |  |
| Stage V | $\geq 0.474$ | $\geq 0.321$ | High risk for mortality |
| Stage IV | [0.149-0.474) | [0.129-0.321) | at-risk (high end) for mortality |
| Stage III | [0.106-0.149) | [0.096-0.129) | at-risk for mortality |
| Stage II | [0.042-0.106) | [0.040-0.096) | at-risk (low end) for mortality |
| Stage I | $< 0.042$ | $< 0.040$ | Low risk for mortality |

### Sensitivity Analysis

We conducted a sensitivity analysis by applying the developed algorithm in the NYC sample and other NYS regions sample, separately. The results showed that the odds ratio of chronic co-morbid conditions varied, but still typically ranged between 1.0-3.0, which was the same as they were in the total sample. The diagnostic accuracy for COVID-19 related fatal outcome was similar in these two sub-samples, with age model reached diagnostic accuracy between 0.77 to 0.80, represented by the area under the ROC curve. Both samples had an overall diagnostic accuracy of 0.85, with the combined demographic variables, clinical presentation/examination, and chronic co-morbid conditions as predictors. (Table 4)

**Table 4.**
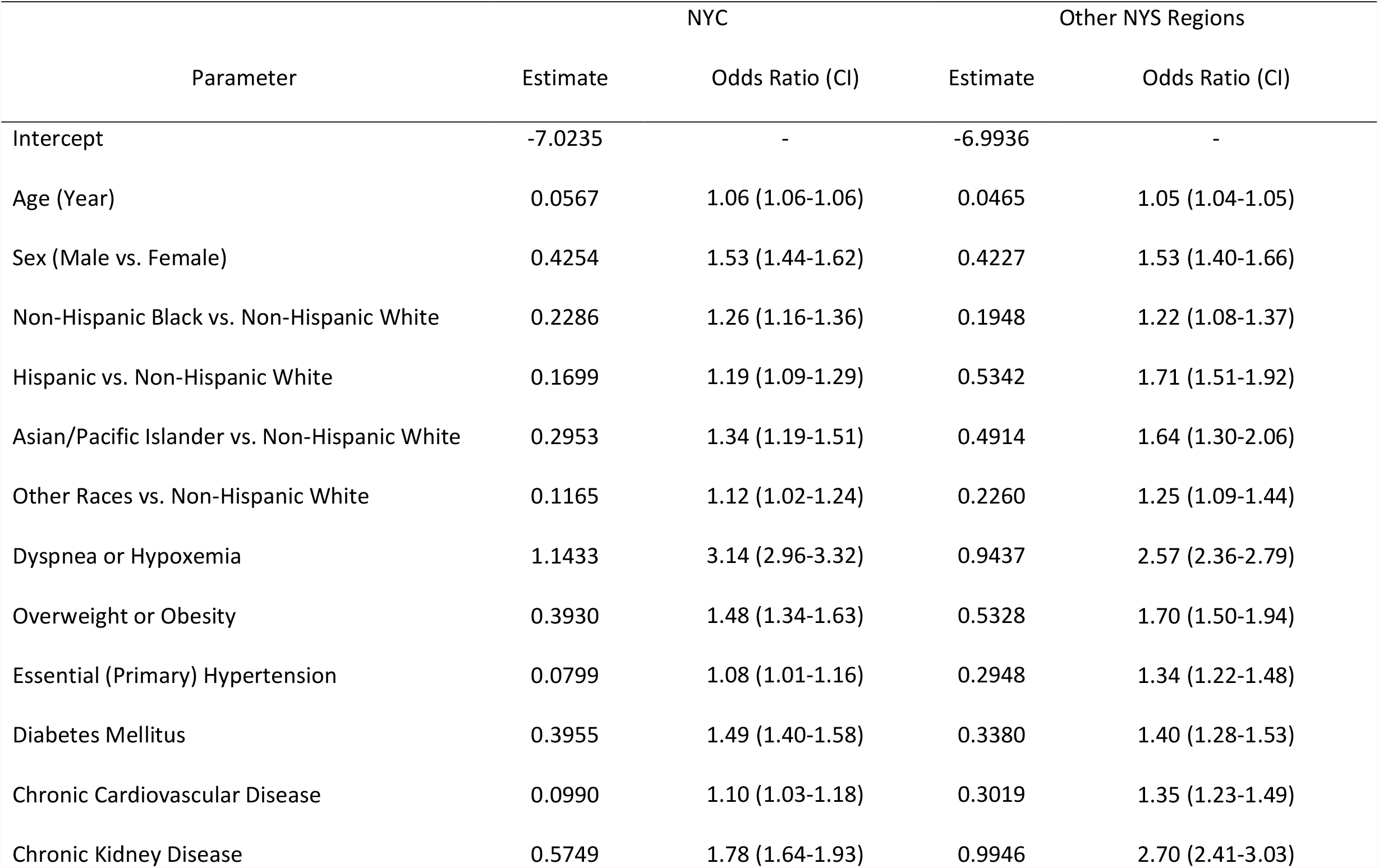
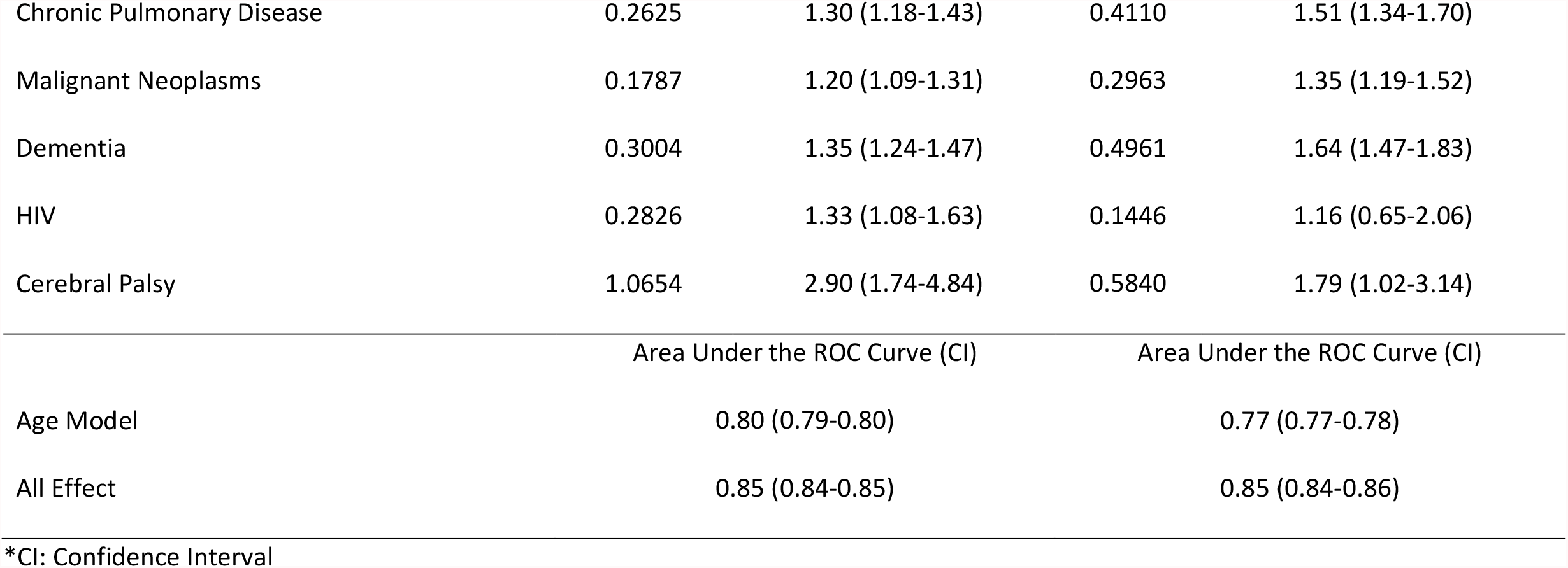
Comparison of Model Estimates for COVID-19 related in-Hospital Death between NYC and Other NYS Regions.

## Discussion

In this study, we evaluated the risk factors for COVID-19 related fatal outcome using a hospital discharge database from NYS. We conducted a manual review and grouping of the secondary diagnoses based on ICD-10-CM codes among the hospital patients, who had a primary diagnosis of COVID-19. Of note, these data were collected relatively early during the severe acute respiratory syndrome coronavirus 2 (SARS-CoV-2) pandemic and before the predominant delta variant emerged. Also, the chronic co-morbid conditions we included in this study were dataset specific. It is difficult for any one dataset to include all the possible co-morbid conditions for COVID-19. Building an algorithm to consider a long list and a very broad spectrum of variables is also challenging, while many of them are inter-related. However, our study nevertheless achieved a similar or even higher diagnostic accuracy as compared to a previous study, which had conducted both the initial investigation and validation for COVID-19 related severe outcome by including a broader spectrum of co-morbid conditions.^6^

Despite this limitation, our study primarily aimed to address three considerations to improve the existing research and knowledge, as well as to explain the scientific thinking and methodology, so that the scientific community can replicate the study in much larger scale datasets with more complete medical information to modify and refine the approach and methodology, and eventually make it useful in clinical practice.

Firstly, the window for early intervention can be short. We need a measurement or scale, which can be applied to rapidly screen the at-risk patients for severe outcome, so that early and timely intervention is possible. In our study, we used patients’ demographic information (age, sex, race/ethnicity), symptom of dyspnea, and medical history of chronic co-morbid conditions, which can be conveniently obtained by inquiry, and hypoxemia, which can be rapidly assessed in out-patient clinic settings.

Secondly, among all the risk factors we discovered in this study for COVID-19 related severe outcome, age may possibly be the most important one, which was consistent with a previous study.^6^ We showed in the study that age by itself can achieve 0.77-0.80 diagnostic accuracy, represented by the area under the ROC curve, while none of the single co-morbid conditions can reach such high diagnostic accuracy in predicting the fatal outcome. Instead, all the multiple co-morbid conditions together only improved around 4% of diagnostic accuracy on top of age. In addition, the odds ratio of the co-morbid conditions typically ranged between 1.0-3.0, after correcting for other demographic variables and dyspnea or hypoxemia; however, the odds of a COVID-19 related fatal outcome increased with much greater magnitude with increased age, such that the odds of 65 year-old patients was 11.9 times that of 18 year-old patients, and 23.6 times that of 5 year-old patients for fatal outcome, after correcting for other demographic variables, dyspnea or hypoxemia, and chronic co-morbid conditions.

These findings do not necessarily mean that the co-morbid conditions weren’t important and should not be considered in decision making for treatment option, but rather indicated that age had greater weight in determining the patients’ fatal outcome. The association between age and many co-morbid conditions have been well established for decades. For predictive purpose, the combined co-morbid conditions without age may also suffice, though age, as a single variable, is likely much easier to manage in clinic. In addition, we treated the co-morbid conditions as dichotomous variables in this study due to the limited information provided in the database. By further stratifying the co-morbid conditions, such as classifying different levels of body mass index (BMI) for overweight/obesity, different levels of hemoglobin A1c for diabetes/hyperglycemia, and different stages and etiologies for chronic kidney disease, etc., there might be more useful information that could potentially be extracted for predictive purpose. It may also be necessary to evaluate in larger samples if the co-morbid conditions matter more among children and young adults than elderly in terms of their associations with the fatal outcome.

The third purpose of this study was to demonstrate the method of cut-off point selection. The current on-going epidemiology studies typically stopped after the predicted probability was calculated. However, predicted probability calculated from such as logistic regression model, was a continuous variable, which didn’t directly reflect the binary outcome, such as mortality status.^7^ In order to use it as a classifier, a cut-off value needs to be chosen. Different predicted probabilities give different sensitivities and specificities in predicting the binary outcomes in the patient sample, which is the fundamental mathematics upon which the ROC curve calculation is based. Although ROC curve gives an overall estimate of diagnostic accuracy, a different cut-off point of odds and/or probability (they can be derived from each other) still gives a different diagnostic accuracy.

In this study, if we were able to achieve an ideal diagnostic accuracy (e.g., sensitivity and specificity above 95%), we would suggest one single cut-off point of odds and/or probability for classification purpose, which should reside on the ROC curve closest to the upper left corner. However, since the diagnostic accuracy was moderate (0.85) in the study, to better facilitate the clinical operation, we proposed two methods for cut-off point selection. For method I, we chose one single cut-off point of odds and/or probability, which was on the ROC curve closest to the upper left corner and classified the patients to two categories, which were high risk vs. low risk for the COVID-19 related fatal outcome (Table 2 & 3). In method II, we proposed a range of cut-off points based on the sensitivity and specificity table (Table 2) and provided a general recommendation to stage the risk to five levels (Table 3). However, physicians can choose a different cut-off point of odds and/or probability based on their clinic needs for patient management.

It is also important to mention that clinical measurements can vary significantly and are often unable to offer a very high-level diagnostic accuracy in predicting the future outcomes. Biomarker development has been going on in other fields for decades to facilitate the clinical outcome predictions and evaluate the pharmaceutical intervention for new drugs, which may be more reliable indicators than the clinical presentation and medical history.^8^ It may be the time to bring this type of work to the field of infectious disease. Further studies are warranted to evaluate laboratory tests and develop laboratory biomarkers in combination with the clinical assessment to improve the diagnostic accuracy and longitudinal prediction, so that at-risk patients can be identified at early stage for intervention with the improved outcome. Hopefully, these advancements will ultimately lead to a reduced mortality rate due to COVID-19 in the general population.

## Data Availability

All data produced in the present work are contained in the manuscript

## Acknowledgement

We greatly appreciate the statistics support and consultation from Ms. Pui Ying Chan and Dr. Sung Woo Lim from Epidemiology Service and data governance and analytical support from Hilary Parton in New York City Department of Health and Mental Hygiene.

## Disclaimer

This publication was produced from raw data purchased from or provided by the New York State Department of Health (NYSDOH). However, the conclusions derived, and views expressed herein are those of the author(s) and do not reflect the conclusions or views of NYSDOH. NYSDOH, its employees, officers, and agents make no representation, warranty or guarantee as to the accuracy, completeness, currency, or suitability of the information provided here. The findings and conclusions also do not necessarily represent the official position of the New York City Department of Health and Mental Hygiene (DOHMH). The authors declared no potential conflicts of interest with respect to the research, authorship, and/or publication of this article.

## References

1. Wong DWL, Klinkhammer BM, Djudjaj S, et al. Multisystemic Cellular Tropism of SARS-CoV-2 in Autopsies of COVID-19 Patients. Cells 2021; 10(8).

2. Center for Disease Control and Prevention. People with Certain Medical Conditions. 2021. https://www.cdc.gov/coronavirus/2019-ncov/need-extra-precautions/people-with-medical-conditions.html (accessed Oct 12 2021).

3. New York State Department of Health. Statewide Planning and Research Cooperative System (SPARCS). 2021. https://www.health.ny.gov/statistics/sparcs/ (accessed Oct 12 2021).

4. Center for Disease Control and Prevention. International Classification of Diseases, Tenth Revision, Clinical Modification (ICD-10-CM). 2020 (accessed Oct 12 2021).

5. Safari S, Baratloo A, Elfil M, Negida A. Evidence Based Emergency Medicine; Part 5 Receiver Operating Curve and Area under the Curve. Emerg (Tehran) 2016; 4(2): 111–3.

6. King JT, Jr., Yoon JS, Rentsch CT, et al. Development and validation of a 30-day mortality index based on pre-existing medical administrative data from 13,323 COVID-19 patients: The Veterans Health Administration COVID-19 (VACO) Index. PLoS One 2020; 15(11): e0241825.

7. Zhang Z. Estimating The Optimal Cutoff Point For Logistic Regression. Open Access Theses & Dissertations. 2018. https://digitalcommons.utep.edu/open_etd/1565 (accessed Nov 29 2021).

8. Huang C, Wahlund LO, Almkvist O, et al. Voxel-and VOI-based analysis of SPECT CBF in relation to clinical and psychological heterogeneity of mild cognitive impairment. Neuroimage 2003; 19(3): 1137–44.

